# Compliance with the Surviving Sepsis Campaign Hour-1 Bundle and impact on patient outcomes in a resource-limited setting: a quality improvement initiative in a referral hospital in Rwanda

**DOI:** 10.1101/2025.08.06.25333121

**Authors:** Jean de Dieu Mahoro, Jean Paul Mvukiyehe, Elisabeth Riviello, Theogene Twagirumugabe

## Abstract

Sepsis and septic shock are common life-threatening conditions associated with high mortality globally. Compliance with the Society of Critical Care Medicine’s Hour-1 Bundle for sepsis management has the potential to improve outcomes, but the bundle is infrequently applied in resource-constrained settings. We assessed level of compliance with the Hour-1 bundle among patients diagnosed with sepsis and septic shock at the Accident and Emergency Department (AED) or Intensive Care Unit (ICU) of University Teaching Hospital of Butare, Rwanda. We conducted training for nurses and doctors in the AED and ICU, with the goal of improving compliance. We re-assessed bundle compliance one month after the training. We used univariate and multivariate analyses to evaluate the association of compliance and fluid resuscitation with hospital mortality while adjusting for severity of illness and other covariates. Our finding showed that seventy-eight patients were enrolled pre-intervention, and 82 post-intervention. The overall compliance with ≥ 4 elements of the Hour-1 bundle before the intervention was 14%, and was 76% after the intervention (p< 0.001). Non-survivors had a significantly lower bundle compliance than survivors (35% vs 80%; adjusted OR 0.1, 95% CI 0.05-0.6; p=0.009). Administered fluids ≥ 2 liters during the first hour was independently associated with higher mortality (adjusted OR 18.0, 95% CI 4.5-71.8; p**<**0.001). These finding suggest that compliance with the Hour-1 sepsis bundle in a resource-constrained setting can be improved with training, and is associated with increased hospital survival. Very liberal fluid resuscitation (≥ 2 liters) in the first hour was independently associated with increased mortality, although bundle-compliant resuscitation was not. Further work is needed, both to implement sepsis bundles in resource-constrained settings, and to ensure that all bundle elements are effective in these settings, and precisely implemented.

## Introduction

Sepsis is a dysregulated host response to infection that is commonly fatal (1). The high mortality associated with sepsis prompted the World Health Organization (WHO) to designate sepsis as a global health priority (2). An estimated 85% of incident sepsis cases worldwide occur in low, lower-middle, and middle-income countries (3). In Rwanda, a low-income country (LIC), sepsis and septic shock are major causes of morbidity and mortality. In a study conducted in the intensive care units (ICUs) of two public referral hospitals in Rwanda in 2016, sepsis was the leading cause of admission to the ICU; of patients enrolled in that study, those suspected or diagnosed with infection within 24 hours of ICU admission had a 2.14 increased odds ratio of death compared to patients without infection (4). Another study conducted in these same public referral hospitals in Rwanda in 2021 demonstrated that mortality associated with sepsis and septic shock was 51.4%, and 82.9% respectively (4). This is much higher than the 38% mortality found in a meta-analysis of studies from North America and Europe among patients diagnosed with septic shock in ICUs (5). This discrepancy in mortality may be due in part to differences in quality of care for septic patients, including early resuscitation, timely administration of appropriate antibiotics, and use of vasopressors (6–9). However, prior trials have demonstrated different responses in high- and low-resource settings to liberal versus conservative fluid resuscitation; outcomes with liberal fluid resuscitation are similar to those with conservative resuscitation in high-income countries (HICs), but outcomes are worse with liberal fluid resuscitation in LICs (6,10,11).

The Society of Critical Care Medicine (SCCM) has published a sepsis Hour-1 Bundle that consists of five components: lactate measurement, blood cultures prior to antibiotic administration, administration of broad-spectrum antibiotics, initiating rapid administration of 30 ml/kg of crystalloid fluid if there is hypotension or lactate>4 mmol/L, and vasopressor initiation if there is hypotension (12–15). In a study in the Netherlands, and another study in Japan, compliance with the bundle was associated with improved hospital survival (16,17). The bundle is a standard practice for sepsis management in many highly-resourced areas of the world, but it is uncommonly used in resource-constrained settings. Little is known about the feasibility of implementing the bundle, or about its potential impact on patient outcomes in resource-constrained settings. Nor is it understood whether bundle-compliant fluid resuscitation is associated with better or worse outcomes. We measured compliance with the bundle at baseline in a Rwandan referral hospital Accident and Emergency Department (AED) and ICU; implemented bundle training for nurses and doctors in the AED and ICU; and then re-assessed compliance four weeks after the training. We also sought to understand the relationship between overall bundle compliance, bundle component compliance, and fluid resuscitation amount, with hospital mortality.

## Materials and Methods

### Design and study participants

We performed a prospective cohort study to assess compliance with the SCCM Hour-1 bundle before and after training of healthcare providers, and to assess for an association between bundle compliance and hospital mortality. The study was conducted in the AED and ICU of the University Teaching Hospital of Butare (CHUB), Rwanda. Patients diagnosed with sepsis by treating clinicians in the AED or ICU were screened for enrollment in the study. During daytime hours in the pre- and post-intervention periods, the Principal Investigator and two research assistants screened patient charts and observed patient care, to locate instances of new diagnoses of sepsis as determined by clinicians in the AED and ICU. Once identified, the study staff further screened the patients for eligibility, by assessing whether they met criteria for sepsis using SCCM recommendations: suspected or confirmed infection; and with evidence of organ dysfunction as evidenced by Modified Early Warning Score (MEWS) ≥ 5, Sequential Organ Failure Assessment (SOFA) score ≥2 or a change of initial SOFA by more than 2 points (18,19) In practice, the team used a checklist with each of these scores, and confirmed eligibility when any of the score cutoffs were met; SOFA was never used for initial inclusion, as laboratory values were not back at the time of screening. Eligible patients or their next of kin were approached to provide a written informed consent.

The study was composed of three phases. Phase I included screening, enrolment, and data collection for septic patients before the intervention. Phase II consisted of a two-month-long period of training of all healthcare providers in the AED and ICU on sepsis management using a local guideline contextualized from the Surviving Sepsis Campaign Guidelines (19), including the Hour-1 sepsis bundle. Phase III consisted of screening, enrolment, and data collection for septic patients after the intervention. Phase III was initiated four weeks after the last training session.

#### Intervention

A local guideline for management of sepsis and septic shock was developed based on the Hour-1 Bundle from SCCM (12,19) All healthcare providers from AED and ICU were trained on it with the following learning objectives:

- Timely recognition of sepsis using the SIRS, SOFA, qSOFA, MEWS, and NEWS scores where applicable, using any of these tools as clinical data is available. The training included handouts with the score elements and cutoffs;
- Adequate fluid resuscitation and monitoring within the first hour of recognition of sepsis;
- Timely identification of patients in shock, and use of vasopressors when fluid resuscitation does not allow achievement of an adequate tissue perfusion (Mean Arterial Pressure (MAP) ≥65 mmHg);
- Early recognition and monitoring of the adequacy of tissue perfusion in patients with sepsis or septic shock by measuring and interpreting serum lactate level;
- Early administration of appropriate antimicrobials, with the goal to obtain relevant cultures prior to antibiotic administration, based on the study site (CHUB) empiric antibiotic guidelines.

Two one-hour training sessions included all anesthesiologists, anesthesia residents, general practitioners, and nurses working in the ICU. An additional two one-hour sessions included all nurses and general practitioners in the AED. The trainings were all led by the Principal Investigator (JDDM), and the training materials were given to clinicians. A summary algorithm was displayed in the department’s physician work stations, nurse work stations, and triage rooms as an ongoing reminder after the training.

### Sample size

We hypothesized that the mortality rate was as high as 82% based on a previous study (4), and we projected an absolute reduction of this mortality by 20% after the training intervention on the SCCM Hour-1 bundle. With a study power of 80% and type-1 error of 0.05, the calculated minimum sample required per group was 78 participants.

#### Data collection

The data collected included patients’ baseline characteristics, compliance with blood sampling and infection source site sampling for culture prior to antibiotic administration, amount of intravenous crystalloids given within one hour of sepsis recognition, administration of appropriate empirical antimicrobial drug administration within one hour of sepsis recognition, administration of vasopressors within the first hour when administered fluids were ineffective to achieve a MAP ≥65 mmHg, and laboratory request for lactate level during the first hour of sepsis recognition. Data collection was done at bedside, and included observation both of chart documentation, as well as observed activities performed by the physicians and nurses.

Variables to complete the MEWS and SOFA scores for each patient were collected; many times data was not fully available in the first hour after sepsis recognition, especially laboratory values. Research assistants returned to the patient’s chart after the initial one-hour period to complete the scores as the data became available. The team also recorded discharge date and vital status at discharge. The Research assistants and PI met weekly to review enrollment numbers and assess data quality.

#### Definitions and Data Analysis

We used a cutoff of >5 for both the MEWS and SOFA scores (20,21); missing data for either of these scores was imputed as normal. Since there was no arterial blood gas (ABG) available at the site, we imputed PaO2/FiO2 from Spo2/ FiO2 (22). We defined “complete” bundle compliance as compliance with at least four of the five elements of the bundle.

Descriptive statistics were used to determine the frequency distribution of all quantitative variables including completion of Hour-1 bundle components and mortality. Comparison between categorical variables was done by using Chi-square or Fisher exact tests. The hospital mortality rate was the primary outcome; we compared mortality between Phase 1 and 3, and also between those with complete bundle compliance and those without. We performed binary univariate analyses to look for associations between baseline characteristics and bundle compliance with mortality. We included variables from this analysis that were associated with mortality (p<0.25) in multivariate logistic regressions; we report adjusted odds ratios (ORs), and corresponding 95% confidence intervals (CIs). A *p-value* < 0.05 was considered statistically significant. All analyses were done with SPSS version 23 IBM, Chicago-IL, USA.

### Ethical considerations

The study was approved by the Institutional Review Board of the University Teaching Hospital of Butare (CHUB) (Ref-REC/UTHB/048/2021). Patients or next of kin provided a written consent prior to their enrollment. The privacy and confidentiality of data were ensured; data were collected from medical records and charts as well as observation at bedside. Hard copies of collected data were stored in a closed cupboard accessible only to the PI and research assistants, and they were entered into Excel by the PI. The dataset was password-protected on the PI’s computer, with a backup on a secure google drive.

## Results

### Enrollment

Phase 1 occurred from December 1, 2021 to August 19, 2022; phase 2 occurred from September 1, 2022 to November 1, 2022; and phase 3 occurred from November 28, 2022 to August 25, 2023. During Phase 1, 81 patients were eligible, and of them, 78 patients consented and were enrolled; three patients died within less than one hour of admission (Fig 1). During Phase 3, 84 patients were eligible, and 82 patients were enrolled; two patients died within the first hour of admission. Of those enrolled, 43 patients were in the AED and 35 in the ICU in phase 1; 38 patients were in the AED and 44 patients in the ICU in phase 3. All eligible patients who survived their first hour after sepsis recognition provided their consent to participate in the study (either directly or via next of kin). All patients designated as septic by clinicians were confirmed to be septic by the research assistants using SCCM criteria.

**Fig 1.**
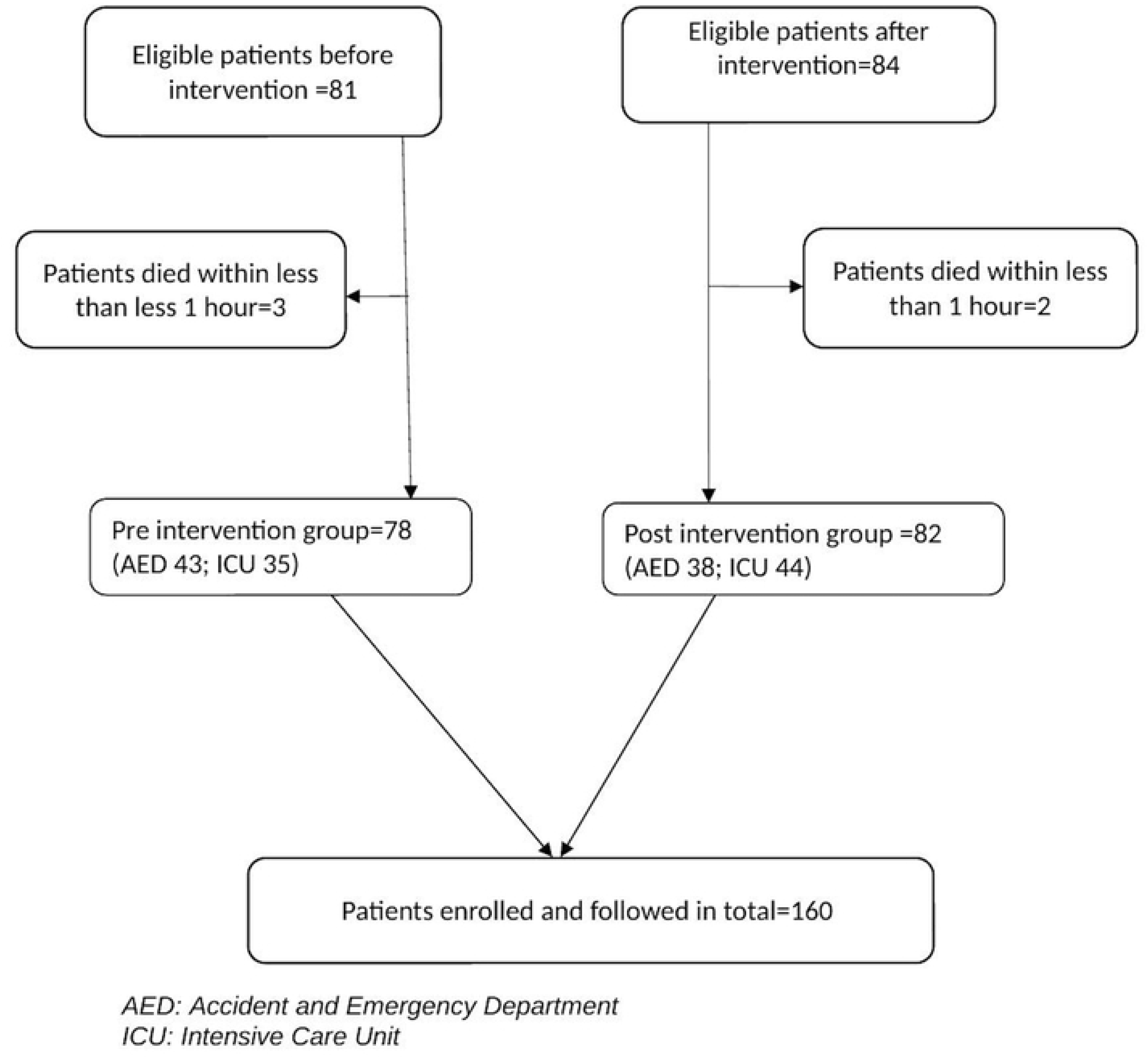
Screening and enrolment

### Pre and post-intervention period baseline patient characteristics

In total, 160 participants were recruited for the study. Of these, 75 (46.9%) were female, and the median age with interquartile range was 41 (30–61) years (Table 1). More than half of participants (84 participants, 52.5%) were admitted due to surgical conditions, and almost all of those (82, 51.2% of the total), had intrabdominal sepsis. The patients in Phase 1 were more severely ill than those in Phase 2: the median (IQR) SOFA score before the intervention was 11 (11–13) compared with 5 (4–10) during the post-intervention period (p<0.001). Similarly, the median (IQR) MEWS before the intervention was 7 (5–9) whereas it was 3 (2-7.2) in the post-intervention period (p<0.001).

**Table 1.** Baseline demographic and clinical characteristics in each period.

| **Variable** |  | ***Before***<br>***intervention***<br>***N=78 (%)*** | ***After***<br>***intervention***<br>***N=82 (%)*** | ***P value*** |
| --- | --- | --- | --- | --- |
| Sex | Female N (%) | 34 (43.5) | 41 (50.0) | 0.417 |
| Age | Median (IQR) | 52 (35.7-65.5) | 44 (30.0-63.2) | 0.108 |
| MEWS>5 | N (%) | 53 (67.9) | 23 (28.0) | <0.001 |
| SOFA>5 | N (%) | 63 (84.0) | 39 (47.6) | <0.001 |
| Surgical vs medical condition | Patient with surgical condition N (%) | 54 (69.2) | 30 (36.6) | <0.001 |
| Source of infection | CNS; N (%) | 7 (9.0) | 18 (22.0) | <0.001 |
|  | Respiratory; N (%) | 11 (14.1) | 33 (40.2) |  |
|  | Abdomen; N (%) | 53 (67.9) | 29 (35.4) |  |
|  | Other; N (%) | 7 (9.0) | 2 (2.4) |  |

*MEWS: Modified Early Warning Score; SOFA: Sequential Organ Failure Assessment; CNS: Central Nervous System. Other includes bacteremia of unknown source and urinary tract infection.*

#### Compliance with Hour-1 bundle in the two periods

Overall, compliance with at least 4 elements of the bundle occurred in 14.1% of cases during the pre-intervention period and 75.6% in post-intervention period (p<0.001) (Table 2, Fig S1). The compliance in measuring lactate before and after the intervention was similar in the two periods (25.6% vs 29.3%, p=0.609). However, the compliance with obtaining culture before antibiotics was significantly improved after the intervention (33.3% vs 90.2%, p<0.001). The administration of appropriate broad-spectrum antibiotics was also much higher in the post-intervention period (29.5% vs 81.7%, p<0.001). The compliance with fluid resuscitation by the guideline was numerically higher in Phase 3 than Phase 1, but it did not reach statistical significance (78.2% vs 89%; p=0.065). The administration of vasopressors if indicated by the clinical conditions was adequate in 52.6% cases vs 86.6% (p<0.001) during the pre-intervention and post-intervention periods respectively.

**Table 2.** Compliance with Hour-1 bundle in the two periods.

| ***Component of the bundle*** | ***Before intervention***<br>***N (%)*** | ***After intervention***<br>***N (%)*** | ***P-value*** |
| --- | --- | --- | --- |
| Measure lactate level | 20 (25.6) | 24 (29.3) | 0.723 |
| Obtain sample for culture before empiric antibiotics | 26 (33.3) | 74 (90.2) | <0.001 |
| Administer broad-spectrum antibiotics | 23 (29.5) | 67 (81.7) | <0.001 |
| Fluid administration if indicated | 61 (78.2) | 73 (89.0) | 0.086 |
| Applying vasopressor if indicated | 41 (52.6) | 71 (86.6) | <0.001 |
| Overall compliance (At least 4 bundle elements completed) | 11 (14.1) | 62 (75.6) | <0.001 |

*Both fluid administration and vasopressor administration were counted as compliant if: 1) they were indicated and given, or 2) they were not indicated and not given.*

### Patient outcomes in the two periods

The overall hospital mortality among 160 study participants was 62.5% (Table S1). Mortality was higher in the pre-intervention period than in the post-intervention period (74.4% vs 51.2%, p=0.003). The median hospital length of stay among survivors was not significantly different between the two periods. It was 6.5 (IQR 3.2-13.2) days and 9.5 (IQR 7.2-12.7) days in the pre and post-intervention periods, respectively p= 0.111 (Table S1).

### Factors associated with mortality

In all patients from Phase 1 and 3, we evaluated the association of sex, age, MEWS, SOFA score, surgical vs medical disease, administered fluids ≥ 2 liters in the first hour, source of infection, compliance with Hour-1 bundle, and compliance with each component of the Hour-1 bundle, with hospital mortality (Table 3). In univariate analyses, MEWS>5, SOFA>5, surgical disease, ≥ 2 liters administered, and non-respiratory source of infection, were all found to be associated with hospital mortality, with p<0.25. In addition, overall bundle compliance and compliance with each element of the bundle were found to be associated with improved survival, with p<0.25. There appears to be a dose-response relationship between compliance with bundle elements and mortality: increasing from 1 to all 5 of the components of the bundle being performed was associated with a stepwise decrease in mortality (Fig 2).

**Fig 2.**
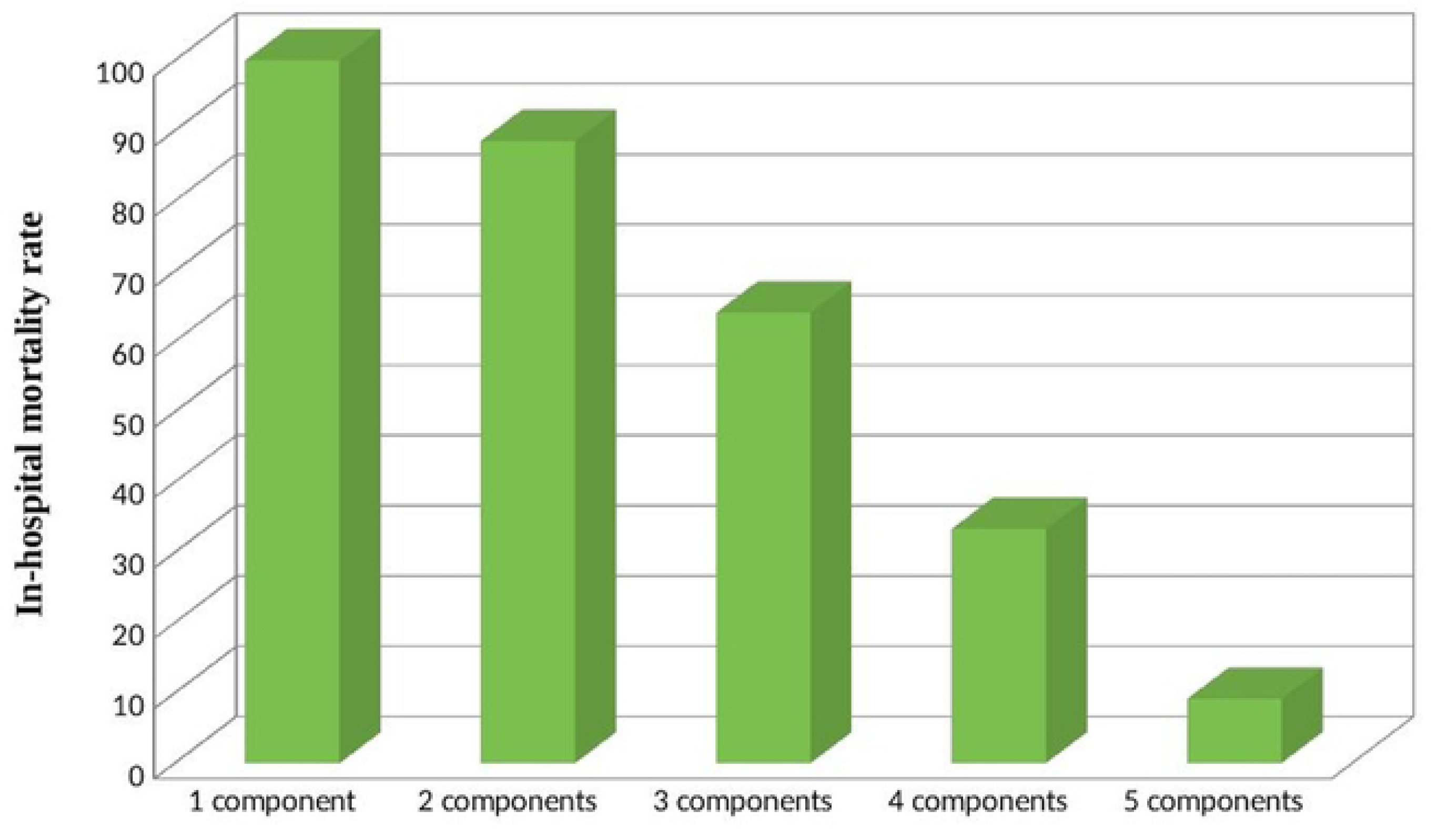
Bundle compliance rate and survival

**Table 3.**
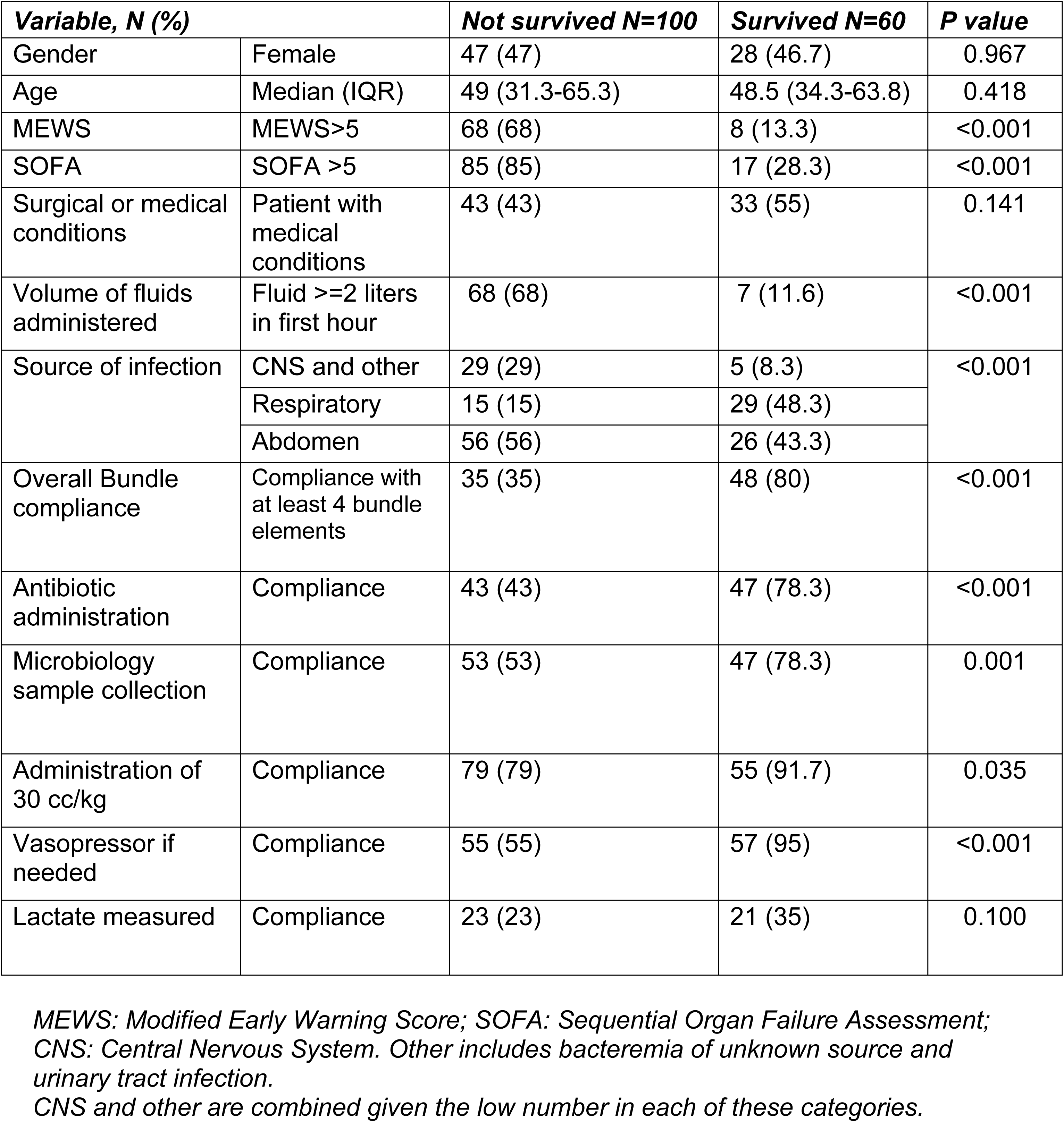
Univariate analysis of factors associated with in-hospital mortality.

We used variables with association with mortality of p<0.25 from the univariate analyses in a multivariate logistic regression analysis. We included overall compliance but not compliance with each component of the bundle, given the overlap of variables. MEWS>5 and administration of IV fluids ≥ 2 liters in the first hour were independently associated with hospital mortality with adjusted odds ratios (AOR): 7.0 (95% CI 1.7-27.2, p=0.005), and 18.0 (95% CI 4.5–71.8, p<0.001) respectively (Table 4). Overall compliance with 4 or more bundle components and having sepsis with a respiratory source were independently associated with reduced hospital mortality (AOR 0.1, 95% CI 0.05-0.6, p = 0.009; AOR 0.02, 95% CI 0.00-0.1, p<0.001 respectively.)

**Table 4.** Multivariate analysis of factors associated with hospital mortality.

| **Variable, N (%)** |  | **Not survived**<br>**N=100** | **Survived**<br>**N=60** | **P value** | **AOR [95%**<br>**CI]** |
| --- | --- | --- | --- | --- | --- |
| MEWS | MEWS $> 5$ | 68 (68) | 8 (13.3) | 0.005 | 7.0 [1.7-27.2] |
| SOFA | SOFA $> 5$ | 85 (85) | 17 (28.3) | 0.083 | 2.7 [0.8-8.7] |
| Surgical or medical conditions | Patient with medical conditions | 43 (43) | 33 (55) | 0.112 | 6.7 [0.6-71.1] |
| Volume of fluids administered | Fluid $\geq 2$ liters in first hour | 68 (68) | 7 (11.6) | $< 0.001$ | 18.0 [4.5-71.8] |
| Source of infection | CNS and other | 29 (29) | 5 (8.3) | Reference |  |
| | Respiratory | 15 (15) | 29 (48.3) | $< 0.001$ | 0.02 [0.00-0.1] |
|  | Abdomen | 56 (56) | 26 (43.3) | 0.749 | 0.6 [0.06-7.3] |
| Overall Bundle compliance | Compliance to at least 4 bundle elements | 35 (35) | 48 (80) | 0.009 | 0.1 [0.05-0.6] |

*MEWS: Modified Early Warning Score; SOFA: Sequential Organ Failure Assessment; CNS: Central Nervous System. Other includes bacteremia of unknown source and urinary tract infection.*
*CNS and other are combined given the low number in each of these categories.*
*AOR: adjusted odds ratio*

The Hosmer and Lemeshow Test of the model confirms that there is no significant difference between the observed and predicted values, as the chi-square value of 7.625 with 8 degrees of freedom has a p-value of 0.471, which is greater than 0.05. This suggests that the model is a good fit for the data (S2 Table).

## Discussion

Almost 49 million incident cases of sepsis occur annually, with 11 million deaths, representing more than 19.7% of all global deaths (3). Incidence of sepsis and sepsis-related mortality remain enormously high in low- and middle-income countries and in Sub-Saharan African countries in particular (23). SCCM’s international guidelines, recently updated under the Surviving Sepsis Campaign, have demonstrated effectiveness in reducing mortality in resource-rich settings (16,17).

Our study demonstrates that training on the Hour-1 sepsis bundle is feasible, and can result in a marked increase in compliance with at least four of five bundle elements. Our adjusted analyses also suggest that increased compliance with the bundle is associated with decreased mortality.

Of note, the component variable of measuring lactate did not statistically improve between periods; this is likely due to inconsistent availability of point of care arterial blood gas analysis with lactate (6).

In addition, bundle-consistent intravenous fluid resuscitation of 30 ml/kg was associated with improved survival in univariate analysis (Table 3), while intravenous fluid resuscitation of 2 liters or more was associated with higher mortality in both univariate and multivariate analyses (Tables 3 and 4). Given the lower weight of many of our patients, 30 ml/kg is often much less than 2 liters. We examined both compliance with 30 ml/kg resuscitation and incidence of ≥ 2 liters resuscitation because previous trials have demonstrated that liberal fluid resuscitation in both adults and children with sepsis in resource-constrained settings is associated with higher mortality (6,11). Our study confirms that very large amounts of fluid may be harmful in LIC settings; however, it also demonstrates that bundle-compliant 30 ml/kg resuscitation may be associated with improved survival in LIC settings. These findings underscore the need for international guidelines that incorporate evidence from trials from a variety of contexts and populations, including resource-constrained settings (24)

To our knowledge, this is the first study to evaluate the impact of compliance with the Hour-1 bundle on the outcome of patients admitted with sepsis in a resource-constrained setting. This is a pragmatic study reflecting real-world circumstances; our results, both in terms of feasibility of improving bundle uptake, and in the suggestion of improved outcomes with improved bundle compliance, are important for other clinicians in similar resource-constrained settings.

Our study has several limitations. First, hospital mortality is our primary outcome, and we do not have data on other important outcomes for patients with critical illness, including longer-term mortality and functional status. Second, there may have been some Hawthorne effect, particularly after the intervention when clinicians knew what elements were being observed; however, using bedside observation gave us very accurate data on bundle compliance in the first hour, which would be impossible to obtain without bedside presence. Third, we have used a before-and-after design, which is at risk for confounding factors and influence of secular trends; however, we used multivariate logistic regression to control for factors most clearly associated with mortality, including illness severity scores; and we are able to directly compare bundle compliance with mortality across all patients, regardless of study phase. Finally, we assessed the post-intervention compliance with the bundle at four weeks from the intervention; we were unable to assess compliance later to understand longer-term retention of learning.

## Conclusion

Compliance with the Hour-1 bundle of the Surviving Sepsis Campaign is low in resource-limited settings. However, it can be markedly increased with a short training; in addition, compliance may improve patient outcomes. Notably, the impact of fluid resuscitation was sensitive to the particular amount administered, with bundle-compliant resuscitation associated with improved outcomes, and more liberal resuscitation associated with worse outcomes. Guidelines and bundles that are validated in a variety of contexts are needed. Our study demonstrates that it is possible to increase compliance with a bundle with relatively few resources, and increased compliance has the potential to have an important positive impact on patient outcomes.

## Data Availability

Data are available upon reasonable request from the corresponding author

N//A

## Acknowledgments

We are thankful to the Directorate of Research at the University Teaching Hospital of Butare for their support. We also thank UMUHIRE Peace and NISHIMWE Delphine for their excellent research assistance.

## Supporting information

S1 Fig. Level of compliance with each component of the Hour-1 bundle and overall, during the two periods

S1 Table. Patient outcomes by period

S2 Table. Hosmer and lemeshow test

## References

1. Singer M, Deutschman CS, Seymour C, Shankar-Hari M, Annane D, Bauer M, et al. The third international consensus definitions for sepsis and septic shock (sepsis-3). Vol. 315, JAMA - Journal of the American Medical Association. American Medical Association; 2016. p. 801–10.

2. Reinhart K, Daniels R, Kissoon N, Machado FR, Schachter RD, Finfer S. Recognizing Sepsis as a Global Health Priority — A WHO Resolution. New England Journal of Medicine. 2017 Aug 3;377(5):414–7.

3. Rudd KE, Johnson SC, Agesa KM, Shackelford KA, Tsoi D, Kievlan DR, et al. Global, regional, and national sepsis incidence and mortality, 1990–2017: analysis for the Global Burden of Disease Study. The Lancet. 2020 Jan 18;395(10219):200–11.

4. Hopkinson DA, Mvukiyehe JP, Jayaraman SP, Syed AA, Dworkin MS, Mucyo W, et al. Sepsis in two hospitals in Rwanda: A retrospective cohort study of presentation, management, outcomes, and predictors of mortality. PLoS One. 2021 May 1;16(5 May 2021).

5. Vincent J louis, Jones G, David S, Olariu E, Cadwell KK. Frequency and mortality of septic shock in Europe and North America : a systematic review and meta-analysis. 2019;1–11.

6. Andrews B, Semler MW, Muchemwa L, Kelly P, Lakhi S, Heimburger DC, et al. Effect of an early resuscitation protocol on in-hospital mortality among adults with sepsis and hypotension: A randomized clinical trial. JAMA - Journal of the American Medical Association. 2017 Oct 3;318(13):1233–40.

7. Kumar A, Ellis P, Arabi Y, Roberts D, Light B, Parrillo JE, et al. Initiation of inappropriate antimicrobial therapy results in a fivefold reduction of survival in human septic shock. Chest. 2009 Nov 1;136(5):1237–48.

8. Kollef MH, Sherman G, Ward S, Fraser VJ. Inadequate antimicrobial treatment of infections: A risk factor for hospital mortality among critically III patients. Chest. 1999;115(2):462–74.

9. Kumar A, Roberts D, Wood KE, Light B, Parrillo JE, Sharma S, et al. Duration of hypotension before initiation of effective antimicrobial therapy is the critical determinant of survival in human septic shock. Crit Care Med. 2006 Jun;34(6):1589–96.

10. Nathan I. Shapiro +Ivor S. Douglas,+Roy G. Brower,. Early Restrictive or Liberal Fluid Management for Sepsis-Induced Hypotension. New England Journal of Medicine [Internet]. 2023 Feb 9;388(6):499–510. Available from: http://www.nejm.org/doi/10.1056/NEJMoa2212663

11. Maitland K, Kiguli S, Opoka RO, Engoru C, Olupot-Olupot P, Akech SO, et al. Mortality after Fluid Bolus in African Children with Severe Infection. New England Journal of Medicine. 2011 Jun 30;364(26):2483–95.

12. Society of Critical Care Medicine. Early Identification of Sepsis on the Hospital Floors: Insights for Implementation of the Hour-1 Bundle. 2019.

13. Rhodes A, Evans LE, Alhazzani W, Levy MM, Antonelli M, Ferrer R, et al. Surviving Sepsis Campaign : International Guidelines for Management of Sepsis and Septic Shock : 2016. Vol. 43, Intensive Care Medicine. Springer Berlin Heidelberg; 2017. 304–377 p.

14. Siddharth Dugar, Chirag Choudhary, Abhijit Duggal. Sepsis and septic shock_ Guideline-based management. Cleve Clin J Med. 2020;

15. Levy MM, Rhodes A, Phillips GS, Townsend SR, Schorr CA, Beale R, et al. Surviving Sepsis Campaign: association between performance metrics and outcomes in a 7.5-year study. Intensive Care Med. 2014 Oct 15;40(11):1623–33.

16. Tromp M, Tjan DHT, Van Zanten ARH, Gielen-Wijffels SEM, Goekoop GJD, Van Den Boogaard M, et al. the effects of implementation of the surviving sepsis Campaign in the netherlands. The journal of Medicine [Internet]. 2011; Available from: www.vmszorg.nl

17. Umemura Y, Abe T, Ogura H, Fujishima S, Kushimoto S, Shiraishi A, et al. Hour-1 bundle adherence was associated with reduction of in-hospital mortality among patients with sepsis in Japan. PLoS One. 2022 Feb 1;17(2).

18. McLymont N, Glover GW. Scoring systems for the characterization of sepsis and associated outcomes. Ann Transl Med. 2016 Dec 1;4(24).

19. Evans L, Rhodes A, Alhazzani W, Antonelli M, Coopersmith CM, French C, et al. Surviving sepsis campaign: international guidelines for management of sepsis and septic shock 2021. Intensive Care Med. 2021 Nov 1;47(11):1181–247.

20. Fayed M, Patel N, Angappan S, Nowak K, Vasconcelos Torres F, Penning DH, et al. Sequential Organ Failure Assessment (SOFA) Score and Mortality Prediction in Patients With Severe Respiratory Distress Secondary to COVID-19. Cureus. 2022 Jul 16;

21. Caramello V, Beux V, de Salve AV, Macciotta A, Ricceri F, Boccuzzi A. Comparison of different prognostic scores for risk stratification in septic patients arriving to the emergency department. Italian Journal of Medicine. 2020 Jun 17;14(2):79–87.

22. Coppola S, Pozzi T, Catozzi G, Monte A, Frascati E, Chiumello D. Clinical Performance of Spo2/Fio2 and Pao2/Fio2 Ratio in Mechanically Ventilated Acute Respiratory Distress Syndrome Patients: A Retrospective Study. Crit Care Med [Internet]. 2025 Mar 3; Available from: http://www.ncbi.nlm.nih.gov/pubmed/40029117

23. Stephen AH, Montoya RL, Aluisio AR. Sepsis and Septic Shock in Low-and Middle-Income Countries. Surg Infect (Larchmt). 2020 Sep 1;21(7):571–8.

24. Riviello ED, Sugira V, Twagirumugabe T. Sepsis research and the poorest of the poor. Lancet Infect Dis [Internet]. 2015;15:501–3. Available from: www.cdc.gov/nchs/data/databriefs/db62.htm

